# Privacy-Enhancing Sequential Learning under Heterogeneous Selection Bias in Multi-Site EHR Data

**DOI:** 10.1101/2025.09.26.25336642

**Authors:** Ritoban Kundu, Xu Shi, Kumar Kshitij Patel, Lucila Ohno-Machado, Maxwell Salvatore, Peter X.K. Song, Bhramar Mukherjee

**Affiliations:** Department of Biostatistics, Epidemiology & Informatics, University of Pennsylvania, Philadelphia, U.S.A.; Department of Biostatistics, University of Michigan, Ann Arbor, U.S.A.; Foundations of Data Science, Yale University, New Haven, U.S.A.; Biomedical Informatics & Data Science, Yale University, New Haven, U.S.A.; School of Medicine, Yale University, New Haven, U.S.A.; Department of Genetics, University of Pennsylvania, Philadelphia, U.S.A.; Department of Biostatistics, Yale University, New Haven, U.S.A.; Department of Chronic Disease Epidemiology, Yale University, New Haven, U.S.A.; Department of Statistics, Yale University, New Haven, U.S.A.

## Abstract

**Objective:** To develop privacy-enhancing statistical methods for estimation of binary disease risk model association parameters across multiple electronic health record (EHR) sites with heterogeneous selection mechanisms, without sharing raw individual-level data. We illustrate their utility through a cross-biobank analysis of smoking and 97 cancer subtypes using data from the NIH All of Us (AOU) and the Michigan Genomics Initiative (MGI).

**Materials and Methods:** Large-scale biobanks often follow heterogeneous recruitment strategies and store data in separate cloud-based platforms, making centralized algorithms infeasible. To address this, we propose two decentralized sequential estimators namely, Sequential Pseudo-likelihood (SPL) and Sequential Augmented Inverse Probability Weighting (SAIPW) that leverage external population-level information to adjust for selection bias, with valid variance estimation. SAIPW additionally protects against misspecification of the selection model using flexible machine learning based auxiliary outcome models. We compare SPL and SAIPW with the existing Sequential Unweighted (SUW) estimator and with centralized and meta learning extensions of IPW and AIPW in simulations under both correctly specified and misspecified selection mechanisms. We apply the methods to harmonized data from MGI (*n* = 50,935) and AOU (*n* = 241,563) to estimate smoking-cancer associations.

**Results:** In simulations, SUW exhibited substantial bias and poor coverage. SPL and SAIPW yielded unbiased estimates with valid coverage probabilities under correct model specification, with SAIPW remaining robust under selection model misspecification. Both approaches showed no notable efficiency loss relative to centralized methods. Meta-learning methods were efficient for large sites but failed in settings with small cohort sizes and rare outcome prevalence. In real-data analysis, strong associations were consistently identified between smoking and cancers of the lung, bladder, and larynx, aligning with established epidemiological evidence.

**Conclusion:** Our framework enables valid, privacy-enhancing inference across EHR cohorts with heterogeneous selection, supporting scalable, decentralized research using real-world data.

## 1 Introduction

### Selection Bias and Privacy Constraints

Electronic Health Records (EHRs) are foundational to clinical care and biomedical research, with adoption exceeding 96% of U.S. hospitals following the 2009 Health Information Technology for Economic and Clinical Health (HITECH) Act (Blumenthal and Tavenner, 2010; Blumenthal, 2011; Adler-Milstein and Jha, 2017). Multi-site EHR analyses offer broader generalizability and greater statistical power. In particular, studying relatively rare conditions, such as complications from invasive procedures, adverse drug events, or associations with rare genetic variants requires integrating data from multiple clinical sites to obtain accurate, generalizable, and reproducible results.

Integrating large biobanks is hindered by two key challenges: heterogeneous recruitment and privacy barriers that preclude centralized analysis. Recruitment strategies vary widely, from the disease-enriched surgical cohorts of the Michigan Genomics Initiative (MGI) (Zawistowski et al., 2023) to the demographically representative national cohort of the NIH All of Us Research Program (AOU) (All Of Us Research Programs Investigators, 2019). These differing mechanisms introduce cohort-specific selection bias (Salvatore et al., 2024), which can lead to biased association estimates if unaddressed (Beesley and Mukherjee, 2022; Kundu et al., 2024a). Simultaneously, data governance policies force initiatives like AOU, MGI or the Million Veteran Program to function as secure enclaves, making it difficult to pool raw individual-level data. These widespread challenges, evident in global efforts like the Global Biobank Meta-analysis Initiative (GBMI) (Zhou et al., 2022) underscore the urgent need for scalable, privacy-enhancing methods that enable valid decentralized inference across heterogeneous EHR cohorts.

### Meta-Analysis

Among the available approaches for estimation under data-sharing constraints, meta-analysis remains a widely used solution. The classical fixed-effects meta-analysis combines site-specific association estimates using inverse-variance weighting (Rice et al., 2018). While effective in many settings, this approach can perform poorly for association parameter estimation in unweighted generalized linear models (Efthimiou, 2018) or IPW-weighted causal estimators (Hu et al., 2024), particularly when individual sites have small sample sizes or rare outcome prevalence, leading to unstable or biased estimates.

### Federated and Sequential Methods

Federated learning provides a privacy-enhancing framework for multi-site analysis by enabling the exchange of summary-level statistics to a central node instead of raw individual-level data across institutions. Likelihood-based federated approaches—such as the surrogate likelihood method by Jordan et al. (2019) and its applications to logistic regression (Duan et al., 2019, 2020, 2022) offer efficient and scalable estimation of global parameters under strict data-sharing constraints. Alternatively, estimating-equation-based decentralized methods, including the renewable estimation framework by Luo and Song (2020) and its extensions (Luo et al., 2024; Hu et al., 2024), are particularly well-suited for settings involving rare outcomes or small sample sizes. However, both classes of methods typically assume that each site’s data are representative of the same target population and overlook differences in site-specific sampling or selection mechanisms. Consequently, they may yield biased estimates when such selection biases are present which is a common challenge in integrating multi-site EHR studies.

### Our Contribution

To simultaneously address the challenges of privacy constraints and selection bias in multi-site EHR studies, we propose a sequential estimation framework. Building on the inverse probability weighting (IPW) and augmented IPW (AIPW) estimating equations developed by Kundu et al. (2024b), we embed these estimators within a renewable estimation paradigm suitable for decentralized environments. Our methodology explicitly accommodates site-specific selection models and nuisance parameters while providing valid variance estimation. By leveraging external probability data representative of the target population, the proposed approach delivers robust, scalable, and privacy-enhancing inference. We evaluate its performance through comprehensive simulation studies and demonstrate its practical utility in a cancer phenome-wide association study (PheWAS) using data from the AOU and the MGI.

## 2 Setup

### 2.1 Problem Setup

We aim to estimate the association between a binary disease indicator *D* and covariates ***Z*** = (*Z*_1_, *Z*_2_, …, *Z*_*p*_) in a target population of size *N*. Specifically, we are interested in the estimation of ***θ***_***Z***_ in the following disease risk model:

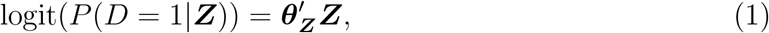

where the covariate vector ***Z*** includes the intercept term. Note that our framework can work for any generalized linear model. Our target population is considered a finite random sample drawn from a hypothetical infinite super-population, similar to the framework considered in Liu et al. (2025). We consider *K* distinct, non-overlapping sites drawn from this target population. Each site *k* (for *k* = 1, 2, …, *K*) is identified by a binary selection indicator, *S*_*k*_. The full set of *p* covariates, ***Z*** (where *p* ≥ 2), is partitioned into an intercept term, a subset ***Z***_1*k*_ where ***Z***_1*k*_ → *D*, ***Z***_1*k*_ *↛ S*_*k*_, and a subset ***Z***_2*k*_ where ***Z***_2*k*_ → *D*, ***Z***_2*k*_ → *S*_*k*_. While this partitioning into ***Z***_1*k*_ and ***Z***_2*k*_ can be specific to each cohort, the overall set of covariates considered, ***Z***_1*k*_ ∪ ***Z***_2*k*_, remains consistent across all sites. Additionally, each cohort *k* may include unique, cohort-specific variables ***W***_*k*_ where ***W***_*k*_ *↛ D*, ***W***_*k*_ → *S*_*k*_. Because disease status *D* can itself influence selection, we define the the probability of selection into site *k* as *P* (*S*_*k*_ = 1|***X***_*k*_) = *π*_*k*_(***X***_*k*_), where ***X***_*k*_ = (*D*, ***Z***_2*k*_, ***W***_*k*_). Thus, each cohort may have its own selection mechanism, potentially differing in both the functional form and the variables ***X***_*k*_ involved.

Moreover, we must adhere to the additional constraint that individual-level raw patient data cannot be shared across different sites, a condition that precludes the use of traditional, centralized statistical methods. To overcome this significant barrier, the subsequent section is dedicated to proposing a novel suite of privacy-enhancing sequential methods.

## 3 Methods

In this section, we present methods for estimating association parameters in the presence of selection bias and under the constraint that individual-level data cannot be shared across sites. We begin by describing the standard unweighted logistic regression, which serves as a baseline as it does not account for selection bias. We then introduce our two proposed approaches: a sequential pseudo-likelihood (SPL) based IPW Logistic Regression, and a doubly robust sequential AIPW (SAIPW) method. For each proposed method, we first detail its theoretical centralized counterpart, the version that could be implemented if all raw data were pooled at a central site. These centralized methods are infeasible for real-world applications and serve as an oracle in our simulations, providing a performance benchmark against which our privacy-enhancing sequential methods are compared.

### 3.1 Unweighted Logistic Regression

#### Centralized Unweighted Logistic Method (CUW)

This method operates under an ideal “centralized” scenario where individual-level data from all participating sites can be stacked together under central node. In this setting, a standard logistic regression using *D* as the binary outcome and ***Z*** as predictors would yield the following score equations for Maximum Likelihood Estimation (MLE). First, the contribution of an individual subject *i* to the score function is defined as:

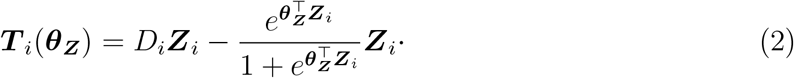

Next, the score function for a specific site *k* is constructed by aggregating the individual contributions of its members:

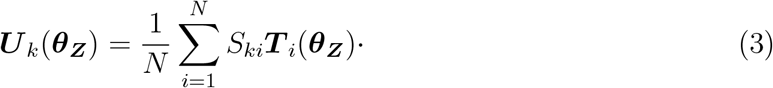

The final Centralized Unweighted (CUW) estimating equation is obtained by summing the score functions across all *K* sites,

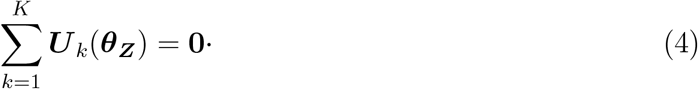

While straightforward to implement, this estimator for ***θ***_***Z***_ can be **substantially biased** when cohort-specific selection mechanisms are present, as demonstrated by Kundu et al. (2024b).

#### Sequential Unweighted Logistic Regression (SUW)

This method leverages the renewable estimation framework developed by Luo and Song (2020), which was originally proposed for unweighted generalized linear models. We adopt their implementation to obtain both parameter estimates and corresponding variance estimates in a sequential paradigm. The SUW approach aggregates summary statistics across sites without incorporating site-specific selection mechanisms, and thus serves as a baseline sequential estimator.

### 3.2 Weighted Logistic Regression

#### 3.2.1 Estimation

##### Centralized Pseudo-likelihood (CPL)

This method also operates under the hypothetical scenario in which individual-level data from all sites can be directly stacked like CUW. To adjust for selection bias, we apply IPW using site-specific selection models. Let ***α***_*k*_ parameterize the selection model for cohort *k*, defined as *π*_*k*_(***X***_*ki*_) = *π*_*k*_(***X***_*ki*_, ***α***_*k*_). The IPW-adjusted score function for the *k*^th^ cohort is:

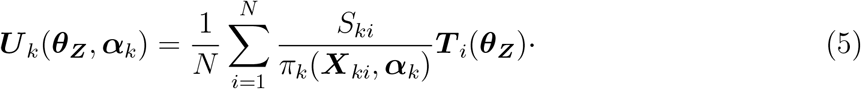

where ***T***_*i*_(***θ***_***Z***_) is defined by equation (2). The selection probabilities *π*_*k*_(*·*) are estimated by integrating an external individual-level probability sample, where *S*_ext_ denotes the indicator variable for selection into the external data. In our application, we use the National Health Interview Survey (NHIS) for this purpose. For each data site *k*, the parameters ***α***_*k*_ are estimated via the Pseudolikelihood (PL) method (Chen et al., 2020), by solving:

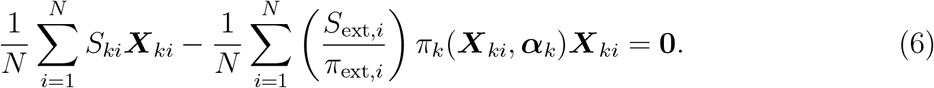

where *π*_ext_ denote the available inclusion probability for each individual in the external probability sample. In the above equation, we specify 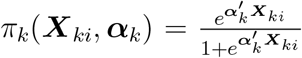. The final CPL estimating equation, aggregating across all cohorts, is given by:

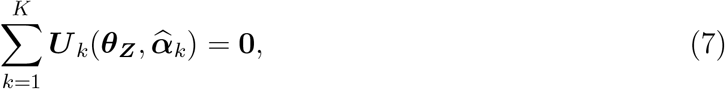

where 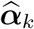 for each site *k* is obtained by solving equation (6). The CPL method yields unbiased estimates when the selection models for all cohorts are correctly specified (Kundu et al., 2024b).

##### Proposal 1: Sequential Pseudolikelihood (SPL)

In this section, we outline the estimation procedure for the privacy enhancing sequential version of the CPL method, designed for settings where raw individual-level data cannot be shared across sites. The SPL approach allows us to iteratively incorporate information from multiple data partners while updating estimates of the association parameters ***θ***_***Z***_. For each site *k* = 1, 2, …, *K*, we define the negative of the Hessian Matrix with respect to parameters ***θ***_***Z***_ as

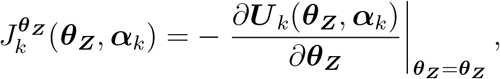

where ***U***_*k*_(*·*) denotes the IPW estimating equation defined in Equation (5).

We begin at site *k* = 1 by estimating the selection parameter 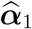 from the PL equation (6). With this estimate, we solve the site-specific estimating equation 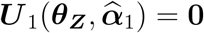 to obtain the initial estimate 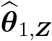. We then compute the corresponding observed information matrix 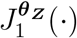 evaluated at 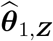 and 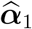.

For each subsequent site *k >* 1, estimation proceeds sequentially. We assume that estimates from the first *k* − 1 sites—namely,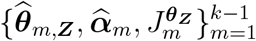—have already been obtained. At site *k*, we begin by estimating the selection model parameters 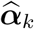 using its local data. Once 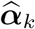 is available, we incorporate it into the following SPL estimating equation:

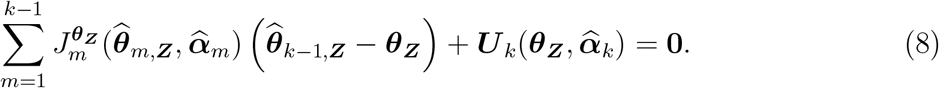

The SPL estimator for the first *k* sites, 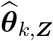, is obtained by solving the estimating equation (8). This equation sequentially combines summary information from the previous (*k* − 1) sites with the score function from the current *k*^th^ site. Figure 1 illustrates this sequential estimation procedure for the case of *K* = 3 data sites, with the nuisance parameters ***η***_*k*_ = ***α***_*k*_.

**Figure 1:**
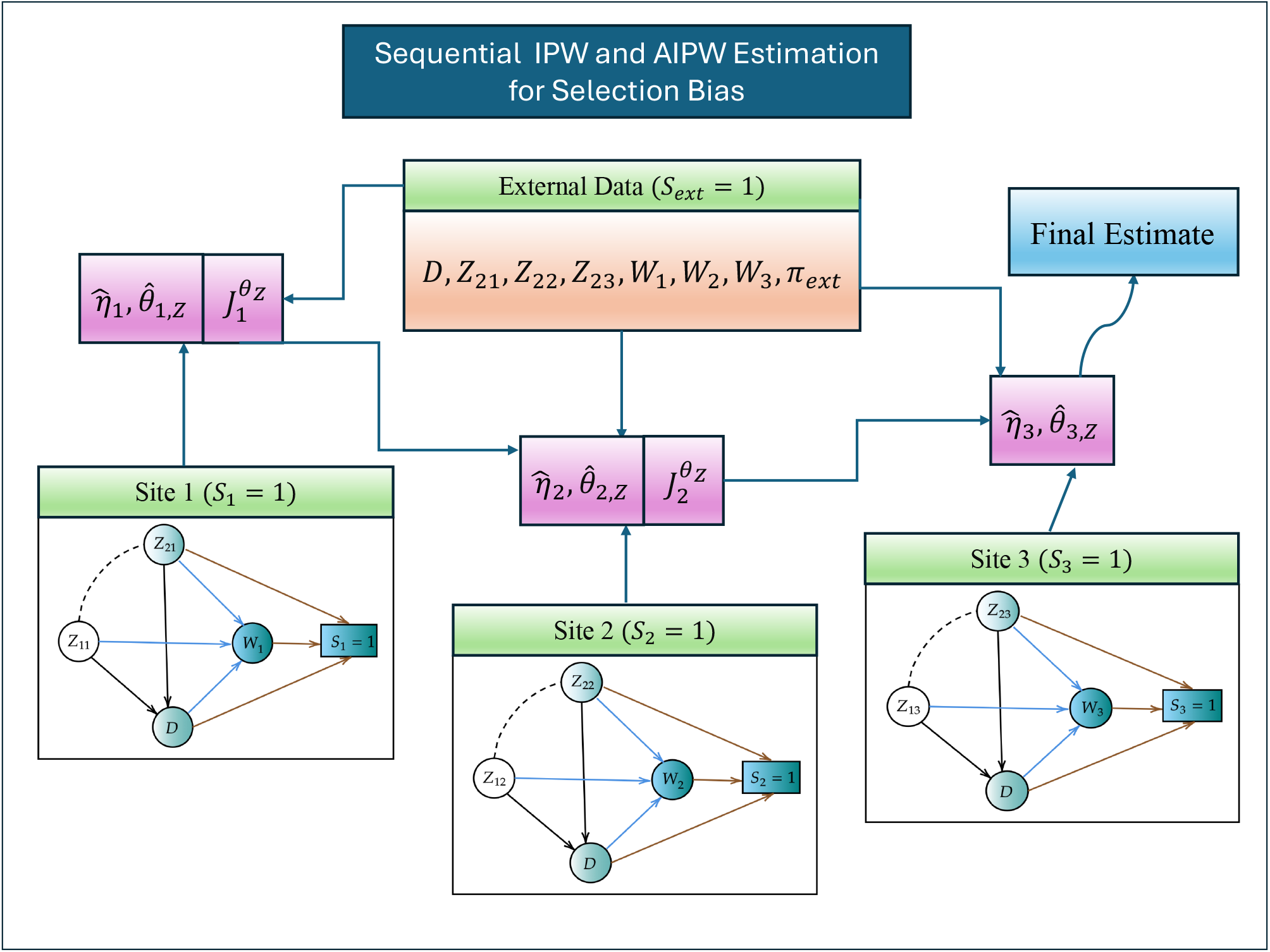
Illustration of the sequential algorithm with *K* = 3 data sites for both the Pseudolikelihood and AIPW methods. Each site sequentially estimates its local nuisance model parameters 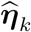 and contributes to the global association parameter update 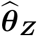 by sharing only summary-level statistics.

#### 3.2.2 Variance Estimation

For CPL estimator, we use the sandwich variance formula from Kundu et al. (2024b). This formula correctly accounts for the additional variance introduced by estimating the nuisance parameters of the selection mechanisms for each site K. The exact form of this estimator is detailed in Supplementary Section S1.1.

We adapt this same variance formula for our proposed SPL estimator. This approach is justified because the SPL estimator converges in probability to the CPL estimator in terms of *ℒ*_2_− norm. The implementation, however, is performed sequentially. For each site *k* = 1, …, *K*, the necessary site-specific components and corresponding terms from external data are computed using the available plug-in estimates. The detailed procedure for this sequential variance computation is presented in Supplementary Section S1.2.

##### Note

Both the CPL and SPL estimators share a critical issue: they produce biased results if the selection model is incorrect for even a single cohort. To address this lack of robustness, we now introduce a doubly robust approach. This method offers protection against model misspecification by leveraging a second, auxiliary score model alongside the selection model. In the following sections, we describe both a centralized and a sequential implementation of this estimator.

### 3.3 Doubly Robust Method

#### 3.3.1 Estimation

##### Centralized Augmented Inverse Probability Weighting (CAIPW)

We adopt the Centralized AIPW method proposed by Kundu et al. (2024b) for multiple cohorts with no overlap in which setting the data can be pooled in a central node. This approach provides double robustness by addressing potential misspecification of the selection propensity score model through augmentation with an auxiliary score function. Specifically, the method combines two key components: (1) a selection propensity score model, and (2) an auxiliary score model. The selection model *π*_*k*_(***X***_*k*_, ***α***_*k*_) for each cohort *k* is estimated via the PL approach described in Equation (6).

The augmented auxiliary score model is constructed by projecting the unweighted logistic score function onto the space spanned by the selection variables ***X***_*k*_. Formally, for cohort *k*, the auxiliary score function is defined as:

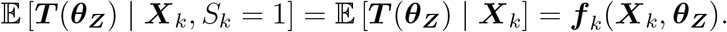

The auxiliary function ***f***_*k*_ (*·*) may be estimated using either parametric or flexible nonparametric methods, depending on the conditional distribution of ***Z***_1*k*_ | ***X***_*k*_. In cohorts where the selection mechanism is independent of the outcome *D*, a more efficient alternative is to define the auxiliary score model using the conditional distribution *D* | ***X***_*k*_. In such cases, the selection variables simplify to ***X***_*k*_ = (***Z***_2*k*_, ***W***_*k*_). In this work, we employ flexible machine learning methods to estimate the nuisance parameter functions. To ensure valid inference, avoid overfitting, and circumvent Donsker conditions, we adopt a cross-fitting strategy following the Double Machine Learning (DML) framework of Chernozhukov et al. (2018).

At each site *k* ∈ {1, 2, …, *K*}, we randomly partition the available data into *L* mutually exclusive folds of equal size, with each fold containing 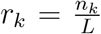 observations, where 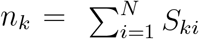 denotes the number of observed individuals in site *k*. Let the combined nuisance parameter be denoted by ***η***_*k*_ = (***α***_*k*_, ***f***_*k*_). For each fold *l* ∈ {1, 2, …, *L*}, we estimate the nuisance functions using the remaining *L* − 1 folds, yielding 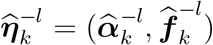. The AIPW estimating equation for site *k* is then defined as:

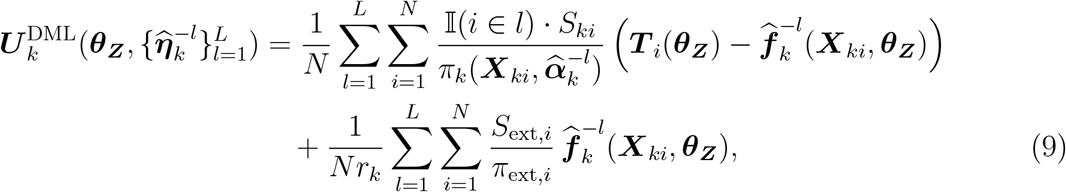

where 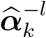 is the estimated selection parameter, and 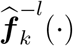 denotes the auxiliary score function trained on the (*L* − 1) training folds for the *L*^th^ split. The final CAIPW estimating equation, aggregating across all cohorts, is then given by:

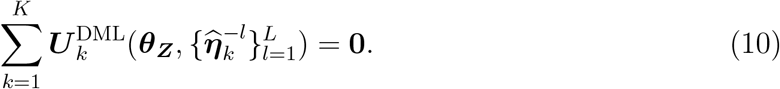

##### Sequential Augmented Inverse Probability Weighting (SAIPW)

We extend the SAIPW method to a sequential framework using the same cross-fitting strategy for estimating nuisance parameters. For each site *k*, we define the gradient of the cross-fitted estimating equation defined in equation (9) with respect to the target parameter as:

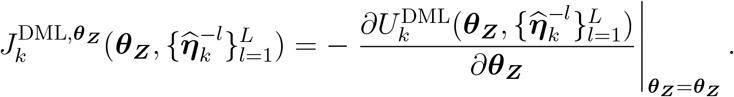

After sequentially computing 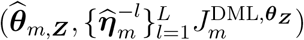 for all *m < k*, we estimate the nuisance parameters 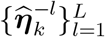 and update ***θ***_***Z***_ at site *k* by solving the following estimating equation:

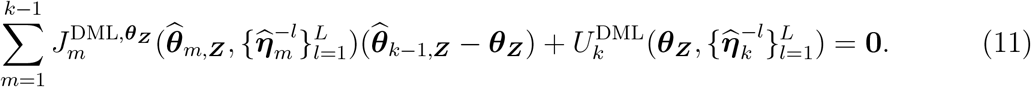

The DML SAIPW estimator for the first *k* sites, 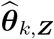, is obtained by solving the estimating equation (11).

#### 3.3.2 Variance Estimation

Under correct specification of both the selection model *π*_*k*_(*·*) and the auxiliary score model ***f***_*k*_ (*·*) for each site *k* ∈ {1, 2, …, *K*}, the influence function corresponding to the DR estimating equation (9) satisfies the Neyman orthogonality property. This property allows us to ignore the contribution of nuisance parameter estimation to the variance, thereby simplifying variance computation. The details of the variance estimation for CAIPW and SAIPW are given in the Supplementary Sections S1.4 and S1.5.

## 4 Simulation

### 4.1 Setup

In this simulation study, we consider a target population of size *N* = 100,000 and generate data from three non-overlapping EHR cohorts. To construct these cohorts, we first simulate three disjoint subpopulations of size *N/*3 each. From these subpopulations, samples are independently drawn to form the individual cohorts, with approximate sample sizes of 14,000, 12,000, and 11,000, respectively. The external probability sample size is 30,000.

We generate *Z*_1_, *Z*_2_, *Z*_3_ from a tri-variate Gaussian distribution. The underlying binary disease outcome *D* is generated according to the logistic regression model:

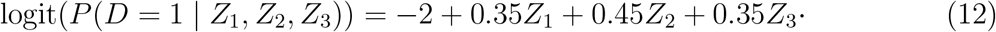

We introduce site-specific selection mechanisms through covariates *W*_1_, *W*_2_, *W*_3_, each of which is generated from a cohort-specific conditional gaussian distribution given *D, Z*_1_, *Z*_2_, *Z*_3_. The selection mechanism for each cohort depends on a distinct subset of covariates: cohort 1 uses ***X***_1_ = (*D, W*_1_, *Z*_2_, *Z*_3_), cohort 2 uses ***X***_2_ = (*D, W*_2_, *Z*_3_), and cohort 3 uses ***X***_3_ = (*D, W*_3_, *Z*_2_).

Hence according the notations in Section 2.1, in cohort 1, the disease model includes ***Z***_11_ = *Z*_1_ and ***Z***_21_ = (*Z*_2_, *Z*_3_), where ***Z***_11_ appears only in the disease model and ***Z***_21_ is shared between the disease and selection models. For cohort 2, ***Z***_12_ = (*Z*_1_, *Z*_2_) and ***Z***_22_ = *Z*_3_, while for cohort 3, ***Z***_13_ = (*Z*_1_, *Z*_3_) and ***Z***_23_ = *Z*_2_. This structure ensures heterogeneity in both covariate availability and selection mechanisms across cohorts to mimic real world settings. The functional forms of the different selection models are provided in Table 1.

**Table 1:**
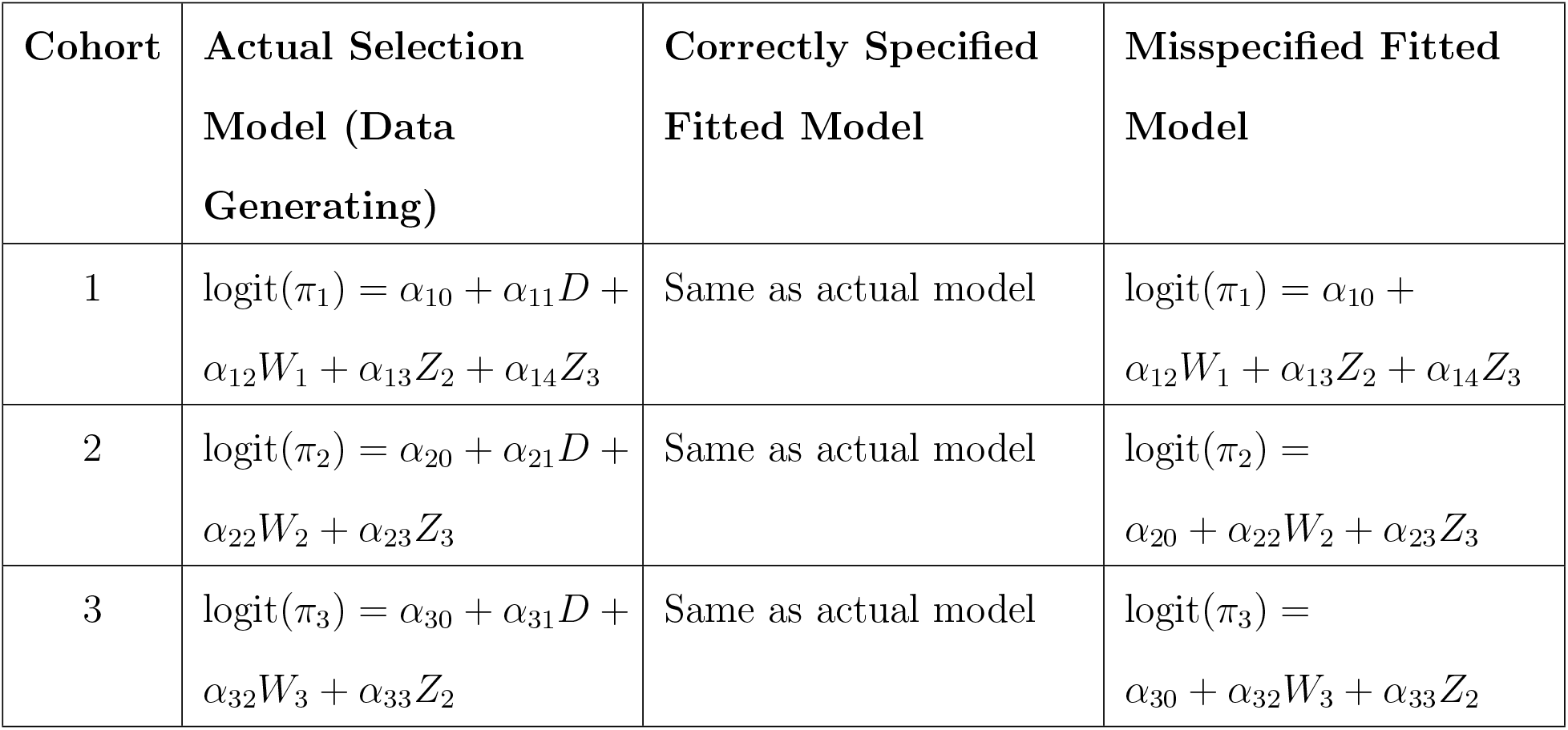
Cohort-specific selection models: Actual data-generating model, correctly specified fitted model, and misspecified fitted model.

### 4.2 Model Fitting and Metrics of Comparison

To evaluate our proposed methods, we compared three distinct analytical approaches for the UW, PL, and AIPW estimators. The first was a centralized analysis, which served as an oracle benchmark assuming all individual-level data could be pooled. The second was our proposed sequential method. For a third comparison, we implemented a classical fixed-effects meta-analysis using the meta package in R (Schwarzer et al., 2007). This provides a standard benchmark method that also relies only on sharing of summary-level data from each site to obtain a pooled estimate.

For the AIPW estimator, we used XGBoost to estimate the auxiliary score functions within a Double Machine Learning (DML) framework with two-fold sample splitting. To assess the robustness of the PL and AIPW methods, we considered scenarios with both correct and incorrect specifications of the selection model. Model misspecification was intentionally induced by omitting the outcome variable *D* from the selection model fitting process in all sites. To evaluate performance, we consider three key metrics. First, the Relative Bias Percentage (RBP) for parameter estimates 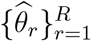 is defined as:

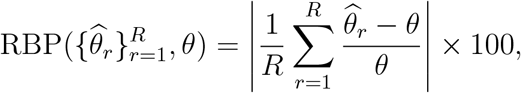

where *R* is the number of simulation replications and *θ* is the true parameter value.

Second, we use the Relative Mean Squared Error (RMSE) to quantify the accuracy of each estimator, defined as the ratio of its MSE to that of the centralized unweighted logistic regression estimator

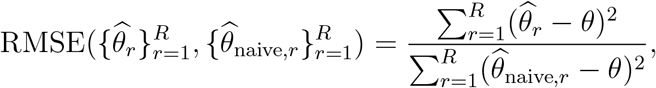

where 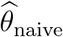 denotes the estimate from the centralized unweighted logistic regression.

Third, to compare the performance of sequential or meta estimators 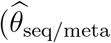 against their centralized counterparts 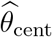, we compute the Relative Efficiency (RE), defined as:

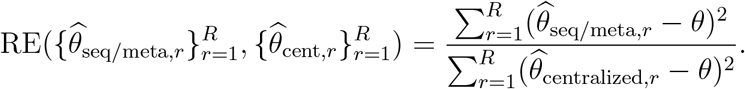

Finally, we evaluate the accuracy of variance estimators using empirical coverage probabilities of the corresponding 95% confidence intervals.

### 4.3 Results

We assess the finite-sample performance of three classes of estimators, centralized (CPL, CAIPW), sequential (SPL, SAIPW) and meta (MPL, MAIPW) under both correctly and incorrectly specified selection models. These are benchmarked against their unweighted counterparts (CUW, SUW, MUW). Results are summarized in Table 2.

**Table 2:**
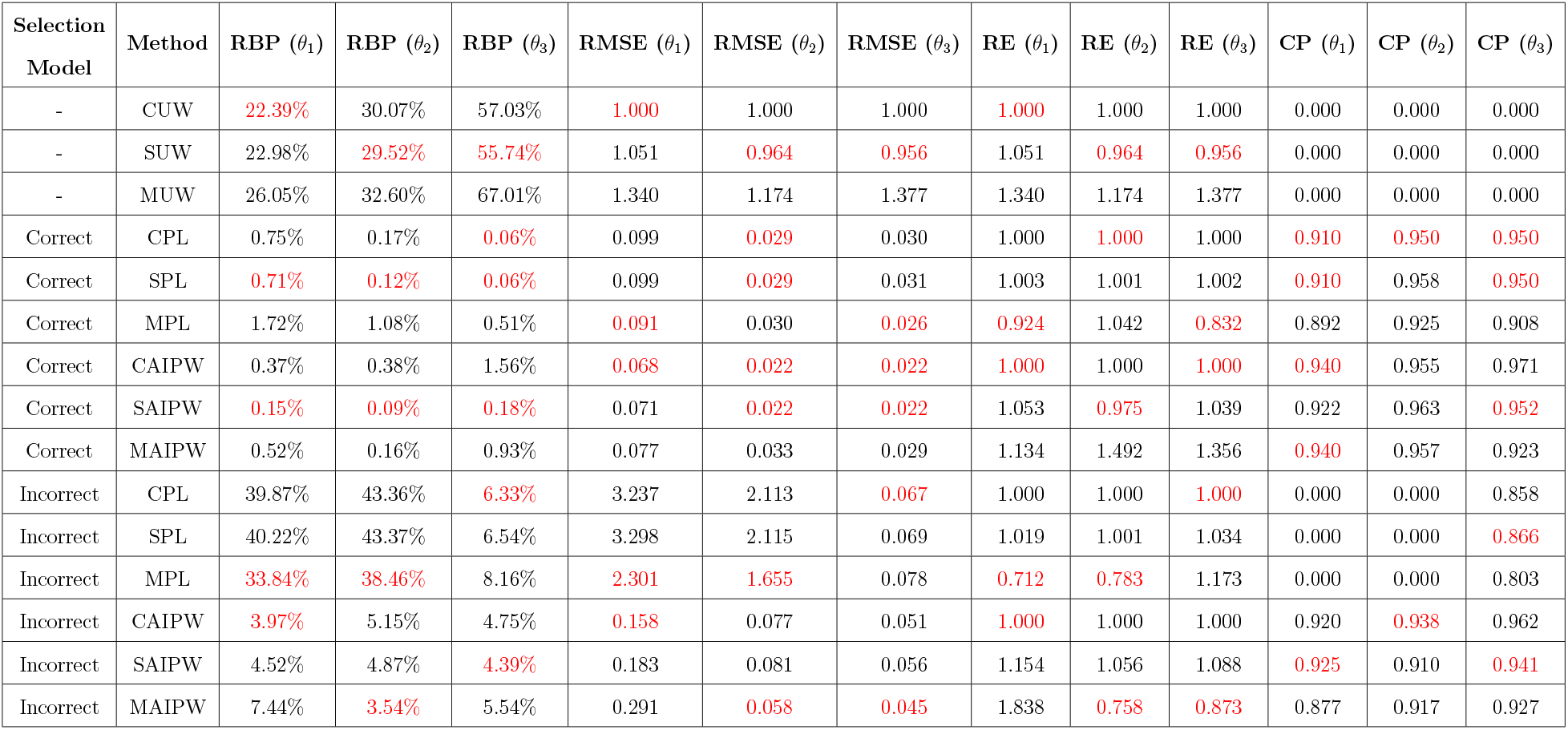
Simulation results comparing centralized (CPL, CAIPW), sequential (SPL, SAIPW), and meta-analysis (MPL, MAIPW) estimators under both correctly specified and misspecified selection models. Metrics reported include relative bias percentage (RBP), root mean squared error (RMSE), relative efficiency (RE), and coverage probability (CP) for each target parameter *θ*_1_, *θ*_2_, and *θ*_3_. The best-performing method for each metric within each modeling scenario and method class is highlighted in red.

| Selection Model | Method | RBP ($\theta_1$) | RBP ($\theta_2$) | RBP ($\theta_3$) | RMSE ($\theta_1$) | RMSE ($\theta_2$) | RMSE ($\theta_3$) | RE ($\theta_1$) | RE ($\theta_2$) | RE ($\theta_3$) | CP ($\theta_1$) | CP ($\theta_2$) | CP ($\theta_3$) |
| --- | --- | --- | --- | --- | --- | --- | --- | --- | --- | --- | --- | --- | --- |
| - | CUW | 22.39% | 30.07% | 57.03% | 1.000 | 1.000 | 1.000 | 1.000 | 1.000 | 1.000 | 0.000 | 0.000 | 0.000 |
| - | SUW | 22.98% | 29.52% | 55.74% | 1.051 | 0.964 | 0.956 | 1.051 | 0.964 | 0.956 | 0.000 | 0.000 | 0.000 |
| - | MUW | 26.05% | 32.60% | 67.01% | 1.340 | 1.174 | 1.377 | 1.340 | 1.174 | 1.377 | 0.000 | 0.000 | 0.000 |
| Correct | CPL | 0.75% | 0.17% | 0.06% | 0.099 | 0.029 | 0.030 | 1.000 | 1.000 | 1.000 | 0.910 | 0.950 | 0.950 |
| Correct | SPL | 0.71% | 0.12% | 0.06% | 0.099 | 0.029 | 0.031 | 1.003 | 1.001 | 1.002 | 0.910 | 0.958 | 0.950 |
| Correct | MPL | 1.72% | 1.08% | 0.51% | 0.091 | 0.030 | 0.026 | 0.924 | 1.042 | 0.832 | 0.892 | 0.925 | 0.908 |
| Correct | CAIPW | 0.37% | 0.38% | 1.56% | 0.068 | 0.022 | 0.022 | 1.000 | 1.000 | 1.000 | 0.940 | 0.955 | 0.971 |
| Correct | SAIPW | 0.15% | 0.09% | 0.18% | 0.071 | 0.022 | 0.022 | 1.053 | 0.975 | 1.039 | 0.922 | 0.963 | 0.952 |
| Correct | MAIPW | 0.52% | 0.16% | 0.93% | 0.077 | 0.033 | 0.029 | 1.134 | 1.492 | 1.356 | 0.940 | 0.957 | 0.923 |
| Incorrect | CPL | 39.87% | 43.36% | 6.33% | 3.237 | 2.113 | 0.067 | 1.000 | 1.000 | 1.000 | 0.000 | 0.000 | 0.858 |
| Incorrect | SPL | 40.22% | 43.37% | 6.54% | 3.298 | 2.115 | 0.069 | 1.019 | 1.001 | 1.034 | 0.000 | 0.000 | 0.866 |
| Incorrect | MPL | 33.84% | 38.46% | 8.16% | 2.301 | 1.655 | 0.078 | 0.712 | 0.783 | 1.173 | 0.000 | 0.000 | 0.803 |
| Incorrect | CAIPW | 3.97% | 5.15% | 4.75% | 0.158 | 0.077 | 0.051 | 1.000 | 1.000 | 1.000 | 0.920 | 0.938 | 0.962 |
| Incorrect | SAIPW | 4.52% | 4.87% | 4.39% | 0.183 | 0.081 | 0.056 | 1.154 | 1.056 | 1.088 | 0.925 | 0.910 | 0.941 |
| Incorrect | MAIPW | 7.44% | 3.54% | 5.54% | 0.291 | 0.058 | 0.045 | 1.838 | 0.758 | 0.873 | 0.877 | 0.917 | 0.927 |

#### Unweighted Methods (CUW, SUW, MUW)

The unweighted methods, which ignore the presence of selection bias, display poor results across all parameters. RBP for *θ*_1_, *θ*_2_, and *θ*_3_ are high, ranging between 22% and 67%. RMSEs for MUW are particularly elevated, with values exceeding 1.3, while CPs are uniformly zero across all unweighted methods.

#### Centralized PL and AIPW Estimators (CPL, CAIPW)

Centralized estimators serve as performance benchmarks. Under correct specification, both CPL and CAIPW yield low bias and RMSEs, with well-calibrated CPs around 0.95. Under incorrect specification, the bias and RMSE of CPL increase noticeably, reflecting its sensitivity to selection model misspecification. In contrast, CAIPW remains robust in such settings, maintaining low bias and RMSE, thereby demonstrating its protection against model misspecification through double robustness.

#### Sequential PL and AIPW Estimators (SPL, SAIPW)

When the selection model is correctly specified, both SPL and SAIPW demonstrate strong performance. SPL achieves very low RBP values (below 1% across all *θ*), with RMSEs closely matching its centralized counterpart (CPL) and CPs near nominal levels. SAIPW, which augments SPL with a flexible auxiliary model, yields even lower RMSEs (e.g., 0.071 for *θ*_1_) and slightly improved coverage. However, under incorrect selection models, SPL’s bias and RMSE increase sharply, particularly for *θ*_1_ and *θ*_2_, where CPs drop to 0, highlighting its sensitivity to mis-specified weights. In contrast, SAIPW remains relatively stable in such settings, with RBPs below 5% and CPs maintained around 0.92-0.94, demonstrating resilience due to its double robustness property. The relative efficiency of all sequential methods remains close to 1, indicating that they do not lose efficiency compared to their centralized counterparts.

#### Meta PL and AIPW Estimators (MPL, MAIPW)

Under these large cohort size scenario, meta estimators, namely MPL and MAIPW, perform comparably to centralized and sequential estimators in terms of bias and RMSE, consistent with Lin and Zeng (2010), which shows that meta-analytic methods can be asymptotically efficient when cohorts share the same target parameter model but differ in nuisance components. Coverage is generally adequate under correct specification; however, MPL shows slight undercoverage for *θ*_1_ (89%), and MAIPW exhibits 88% coverage under selection model misspecification. This undercoverage may stem from the fixed-effect meta-analysis variance formula assuming independence across cohorts, an assumption violated here due to shared external data used in selection weights estimation.

### 4.4 Additional Simulations : Large *K*

This simulation setup is designed to evaluate the performance of CPL, SPL, and MPL in scenarios involving a large number of cohorts with small individual sample sizes and rare disease prevalence. We also include CUW, SUW, and MUW as benchmark methods.

#### Setup

We considered *K* = 20 cohorts, with cohort-specific sample sizes ranging from 230 to 342 and disease prevalence varying between 3.4% and 9.7%. The population disease model followed equation (12), and an external probability sample of size 2000 was used. Site-specific selection covariates *W*_1_, …, *W*_20_, were generated from a cohort-specific Gaussian distribution conditional on *D, Z*_1_, *Z*_2_, and *Z*_3_. For each cohort *k, S*_*k*_ depended on *D, Z*_2_, *Z*_3_, and the site-specific variable *W*_*k*_. All methods were evaluated under correctly specified selection models.

##### Results

From Supplementary Table S1, we observe that for *θ*_1_, the unweighted methods exhibited moderate bias (approximately 15%), likely because *Z*_1_ was not involved in the selection mechanisms. However, for *θ*_2_ and *θ*_3_, relative biases exceeded 80% across all unweighted methods. MPL also showed large biases across all parameters, similar to unweighted approaches. The relative efficiency of MPL compared to CPL reached 3.49 for *θ*_3_, indicating inflated variance. In contrast, SPL and CPL showed comparable performance, with low bias and RMSE across all parameters. For *θ*_2_ and *θ*_3_, both methods outperformed all others, achieving coverage probabilities close to 95%. These findings suggest that even in settings with rare disease prevalence and limited individual-level data, SPL achieves estimation accuracy comparable to CPL, with no meaningful loss of efficiency.

## 5 Cancer PheWAS Application: Association Between Smoking and Cancer Types

### 5.1 Introduction to Data

#### Setup and Goal

We conduct a phenome-wide association study (PheWAS) to investigate the relationship between smoking and a wide range of cancer types using two major EHR datasets: MGI and the NIH AOU Research Program. Smoking is a known risk factor for various cancers, including those of the blood (e.g., acute myeloid leukemia), bladder, cervix, colon, esophagus, kidney, larynx, liver, lung, mouth, pancreas, pharynx, stomach, and trachea (CDC Smoking-Cancer). Our goal is to empirically evaluate these associations using rich EHR-linked cohorts while addressing selection bias and sampling differences across data sources.

#### Data Sources

The AOU program, launched by NIH in 2018, aims to recruit over one million adults across the U.S. with intentional oversampling of underrepresented populations (All Of Us Research Programs Investigators, 2019). We use Version 8 of the Curated Data Repository (CDR V8), with data up to October 1, 2023. After filtering for complete information on smoking, cancer diagnoses, and other covariates, the analytic AOU cohort includes 241,563 individuals. The MGI cohort, initiated in 2012 at Michigan Medicine, recruits participants mainly from preoperative anesthesia visits (Zawistowski et al., 2023), resulting in a clinically enriched cohort ideal for PheWAS. After removing missing data, the final MGI sample includes 50,935 individuals (as of August 2022). For population-level selection adjustment, we use the 2019 National Health Interview Survey (NHIS), a nationally representative survey of U.S. adults conducted by the National Center for Health Statistics. After restricting to complete cases, the NHIS sample comprises 27,152 individuals.

### 5.2 Data Summary

Table 3 presents summary statistics for the MGI, AOU and NHIS (weighted to the U.S. population).

**Table 3:** Summary statistics of key demographic and clinical variables across the MGI (Michigan Genomics Initiative), AOU (All of Us Research Program), and NHIS (National Health Interview Survey) cohorts. Reported values include sample sizes, means and standard deviations for continuous variables, and percentages for categorical variables. NHIS values are weighted using survey weights to reflect population-level estimates. Here, CHD stands for Coronary Heart Disease.

| **Variables** | **MGI** | **AOU** | **NHIS (Weighted)** |
| --- | --- | --- | --- |
| **Sample Size** | 50935 | 241563 | 27152 |
| **Age** | Mean : 58.98<br>Sd : 16.59 | Mean : 55.04<br>Sd : 17.18 | Mean : 47.97<br>Sd: 18.25 |
| **Biological Sex** | Females : 53.07% | Female : 61.93% | Females : 50.89% |
| **Race** | NHW : 89.13% | NHW : 63.87% | NHW: 66.05% |
| **Overall Cancer** | Yes : 52.36% | Yes : 23.35% | Yes : 9.64% |
| **Smokers** | Ever Smoked : 49.28% | Ever Smoked : 39.87% | Ever Smoked: 36.69% |
| **Type 2 Diabetes** | Yes: 30.85% | Yes : 22.72% | Yes : 8.93% |
| **CHD** | Yes : 17.18% | Yes : 14.26% | Yes : 4.61% |
| **BMI** | Mean : 29.86<br>Sd : 7.06 | Mean : 30.08<br>Sd : 7.77 | Mean : 27.85<br>Sd : 5.48 |
| **Sexual Orientation** | NA | Not Straight : 11.17% | Not Straight : 3.10% |
| **Health Insurance** | NA | Yes : 96.01% | Yes: 88.87% |
| **Income** | NA | Over 75K : 39.14% | Over 75K : 45.00% |
| **Region** | NA | West : 24.79%<br>South : 17.14%<br>Northeast : 30.10%<br>Midwest : 21.43% | West : 23.13%<br>South: 37.58%<br>Northeast: 17.85%<br>Midwest : 21.43% |

#### Cancer Data Summary

We began with 100 cancer-related phecodes for our phenome-wide association analysis. The full list of phecodes is provided in Supplementary Section S1 for reference. However, we excluded three phecodes—”149.5”, “196”, and “197”—due to an absence of corresponding cases in the AOU dataset. This left us with 97 distinct cancer types for subsequent analysis. In the AOU cohort, the overall prevalence of any cancer diagnosis was 23.35%. Prevalence estimates for individual cancer types ranged from 0.02% for phecode “149.3” (indicating a rare malignancy of the pharynx) to 6.44% for phecode “195” (representing cancer, unspecified site). In contrast, the MGI cohort exhibited a substantially higher overall cancer prevalence of 52.36%, reflective of its clinically enriched recruitment design. In this cohort, prevalence ranged from 0.06% for phecode “204.3” (representing chronic lymphocytic leukemia) to 38.14% for phecode “195”.

#### Summary of Cohort Characteristics

According to Table 3, MGI participants were older (mean age: 58.98 years) than those in AOU (55.04 years), while NHIS participants were younger (47.97 years). MGI and AOU had a higher proportion of females (53.07% and 61.93%, respectively) compared to NHIS (50.89%). Non-Hispanic Whites were most prevalent in MGI (89.13%), followed by AOU (63.87%) and NHIS (66.05%). Ever-smoking rates were 49.28% in MGI and 39.87% in AOU, compared to 36.69% in NHIS. For comorbidities, MGI showed the highest prevalence of type 2 diabetes (30.85%) and coronary heart disease (CHD; 17.18%), followed by AOU (18.76% and 7.72%), with NHIS lowest (10.32% and 4.53%). Mean BMI was similar in MGI (30.01 kg/m^2^) and AOU (30.15 kg/m^2^), but lower in NHIS (27.85 kg/m^2^). Sociodemographic data available in AOU and NHIS show that AOU had more sexual minorities (11.17% vs. 3.10%), higher insurance coverage (96.01% vs. 88.87%), and fewer participants from the South (17.14% vs. 37.58%) compared to NHIS.

### 5.3 Model Fitting Characteristics

#### Cancer PheWAS Model

For each of the 97 cancer phecodes, we estimated the conditional association between smoking and cancer risk using logistic regression, targeting the U.S. adult population adjusted for biological sex, age and race. Let *D*_*c*_ ∈ {0, 1} denote the binary indicator for the presence of cancer type *c* ∈ {1, 2, …, 97}. For each cancer type, we aim to estimate the target parameter *β*_1_ in the following model:

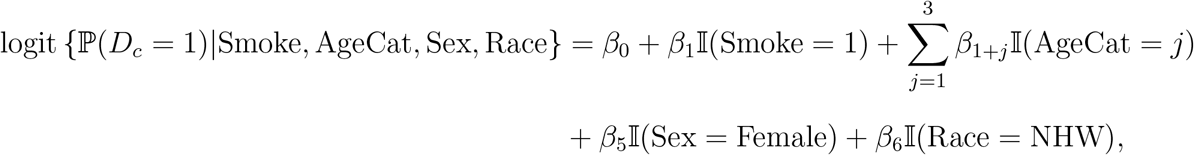

where *j* = 1, 2, 3 denotes age categories of the categorized age variable, namely “AgeVar” corresponding to 35–49, 50–64, and 65+ years; reference: 18–34 years).

#### Selection Models

##### AOU

For the AOU cohort, the selection model incorporated age, biological sex, race, presence of health insurance (HI), sexual orientation (SO; non-straight vs. straight), annual household income above $75,000 (IN), and geographic region (Northeast, South, Midwest, West). We assume the following selection model:

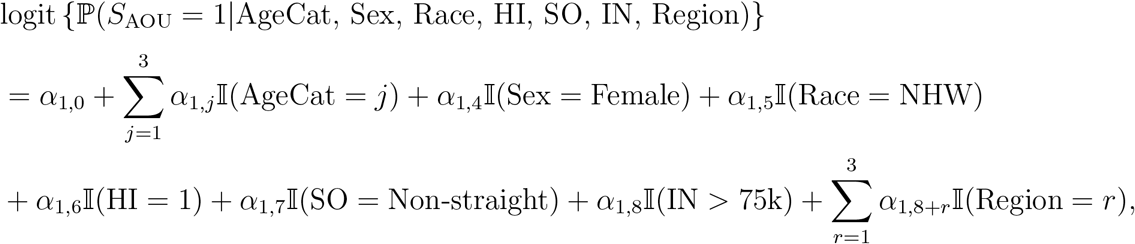

where Region = *r* denotes Northeast, Midwest, and South, with West as the reference category.

##### MGI

The selection model for the MGI cohort included the following variables: age, coronary artery disease (CAD), type 2 diabetes (DI), race (Non-Hispanic White, NHW), overall cancer (C), and smoking. The model is specified as:

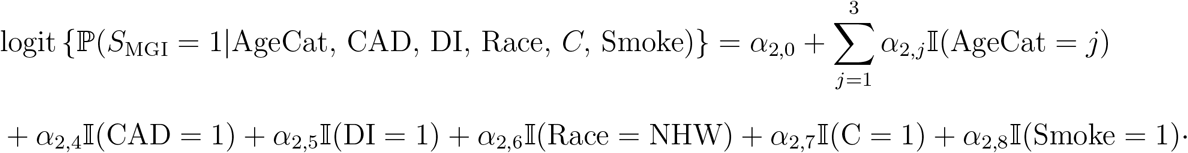

#### Auxiliary Models

This section describes the auxiliary score models used exclusively for the SAIPW method.

##### AOU

In the AOU cohort, the specific cancer subtype does not influence the selection mechanism. Therefore, we construct the auxiliary score models based on the conditional distribution of each cancer subtype given the observed covariates. These include age, biological sex, race, health insurance status, sexual orientation, annual household income, geographic region, and smoking status. The auxiliary score model for cancer subtype *c* depends on the following probability:

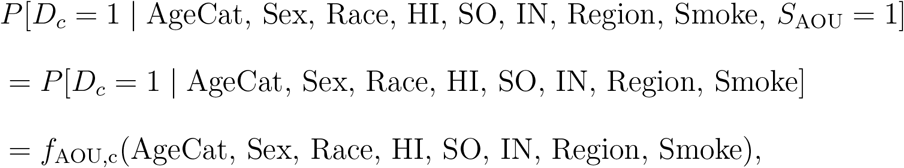

##### MGI

The MGI cohort presents a more complex scenario because cancer subtype is part of the selection mechanism. However, the external reference dataset lacks information on individual subtypes. Since the auxiliary score model conditions on the overall cancer indicator *C*, which is available in the external data, the specific subtype *D*_*c*_ is independent of selection indicator given *C*. Accordingly, the auxiliary score model for cancer subtype *c* depends on the following probability:

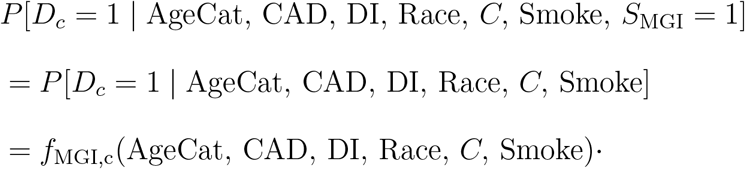

### 5.4 Comparison of Methods and Interpretation of Results

In this analysis we investigated the covariate-adjusted associations between smoking and 97 distinct cancer types using SUW, SPL and SAIPW. All models adjusted for age, sex, and race, and estimated conditional effects of smoking. Therefore, these findings may not align perfectly with marginal real-world association estimates but are intended to reflect more nuanced, adjusted associations that account for potential confounding. Figure 2 illustrates the results obtained from this Cancer PheWAS association study.

**Figure 2:**
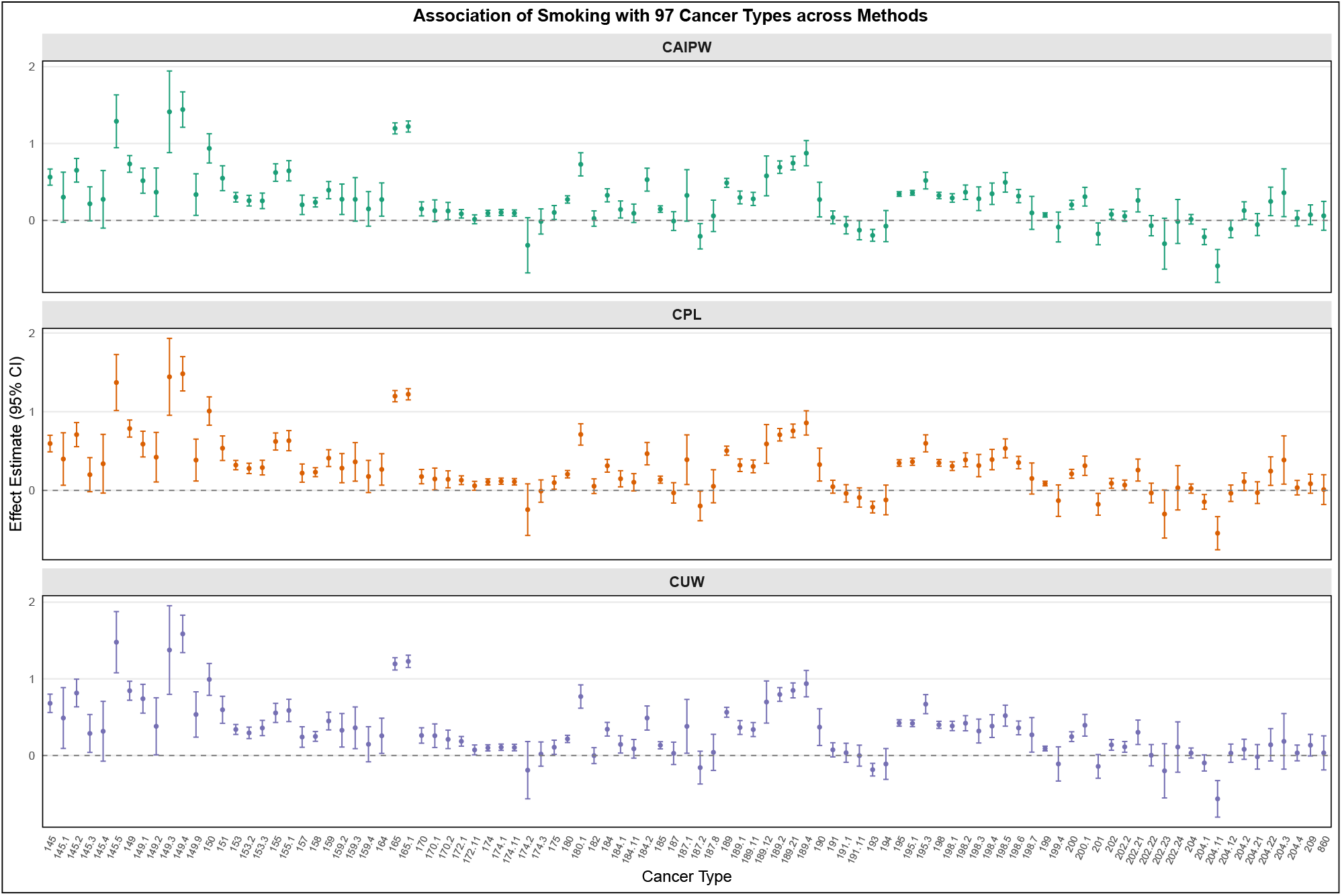
Association estimates between smoking and 97 cancer types using three different methods: SUW (Sequential Unweighted), SPL (Sequential Pseudolikelihood), and SAIPW (Sequential Augmented Inverse Probability Weighting). Point estimates and 95% confidence intervals are shown for each method. The x-axis denotes cancer types and the y-axis represents the effect estimates.

Across cancer types with well-established links to smoking-such as lung, laryngeal, and bladder cancers-both SPL and SAIPW methods yielded consistent positive associations. For example, phecode 145.2 (laryngeal cancer) showed strong estimates from all methods (SUW: 0.82 [0.63, 1.00], SPL: 0.84 [0.66, 1.02], SAIPW: 0.65 [0.50, 0.81]), aligning with clinical evidence that identifies smoking as a major risk factor. Similarly, for lung cancer (phecode 165.1), SAIPW estimated a log-odds of 1.22 [1.15, 1.29], consistent with known high risks associated with smoking.

SUW often produced wider confidence intervals, particularly for rarer cancers, because it does not incorporate information beyond the internal cohort and thus has limited precision. In contrast, SPL and SAIPW provided more stable estimates by integrating site-specific covariates with external reference data. For example, in tonsillar cancer (Phecode 149.2), SUW yielded a 95% confidence interval of [0.01, 0.75], while SPL and SAIPW produced tighter intervals of [0.11, 0.74] and [0.05, 0.68], respectively. This pattern of improved precision under SPL and SAIPW is consistent across cancer types (Figure 3), especially where prevalence is extremely low. Importantly, this improvement does not stem primarily from correcting selection bias, but rather from the fact that SPL and SAIPW borrow additional information from external sources. By leveraging such external data, these methods effectively increase the sample size for rare cancers, thereby enhancing estimation accuracy and reducing variability. While the reported estimates reflect adjusted conditional associations rather than marginal risks, they nonetheless provide important insights into generalizable associations between smoking and cancer across heterogeneous, real-world EHR cohorts.

**Figure 3:**
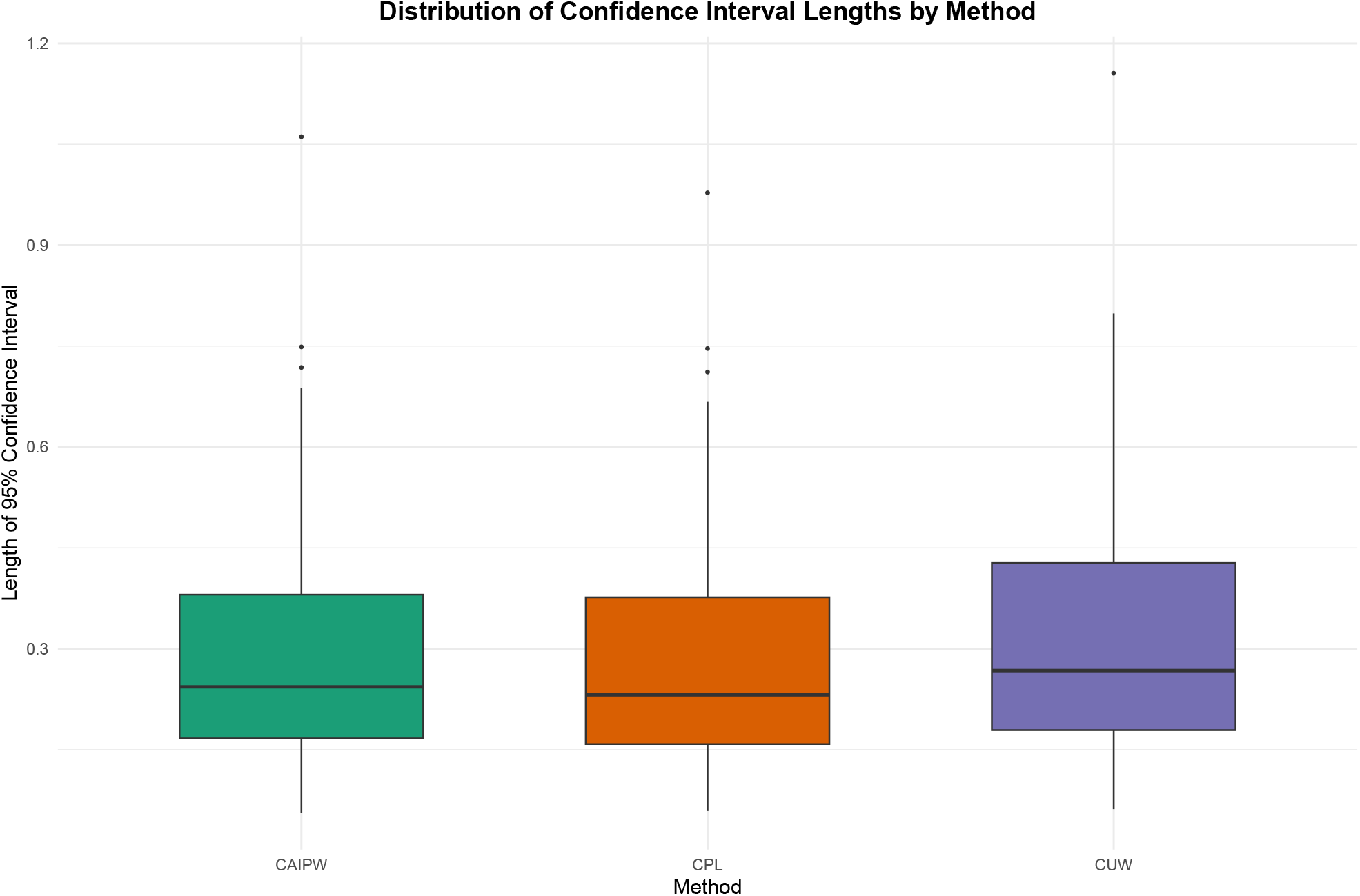
Distribution of 95% confidence interval lengths across methods (SUW, SPL, SAIPW) for all cancer types.

## 6 Discussion and Conclusion

This work introduces a suite of sequential estimation methods for binary outcome regression in multi-site EHR studies where raw individual-level data cannot be shared across sites. We proposed two sequential estimators, namely SPL, and SAIPW that leverage external probability samples to adjust for site-specific selection mechanisms while enhancing patient privacy.

Through extensive simulations, we demonstrated that SPL and SAIPW substantially reduce bias and improve estimation efficiency relative to the naive SUW method, especially under heterogeneous sampling mechanisms and partial covariate overlap. Both the methods had comparable efficiency respect to the hypothetical centralized counterparts. SAIPW, in particular, maintained strong performance under selection model misspecification, benefiting from its doubly robust design. Importantly, the sequential estimators performed comparably to their centralized counterparts, despite operating under stricter privacy constraints.

We also evaluated fixed-effects meta-analysis approaches (MPL and MAIPW), which aggregate site-specific estimates using inverse-variance weighting. Under correct model specification and moderate-to-large cohort sizes, these meta estimators exhibited similar bias and RMSE to sequential and centralized estimators, consistent with theoretical expectations. However, in settings with smaller sample sizes and large *K*, particularly under rare disease prevalence, their performance deteriorated—showing increased bias and variability.

We applied our methods to a PheWAS of smoking and 97 cancer types using two EHR cohorts (AOU and MGI), with NHIS as the reference population. Naive SUW estimates were unstable and showed inflated variability, while SPL and SAIPW produced more stable and interpretable associations. Elevated risks for lung, larynx, bladder, and oral cancers were consistently identified, aligning with established epidemiological evidence.

It’s important to clarify that while our proposed estimators are termed “sequential” the final estimates for both SPL and SAIPW are invariant to the order in which the cohorts are processed. The “sequential” nature of the framework refers to the computational approach used to preserve privacy—analyzing sites one by one—rather than any required ordering of the sites themselves. This order-invariance is supported both theoretically and empirically. For instance, in our simulations with three cohorts, we confirmed that the results were identical across all six possible permutations of the cohort sequence. For practical implementation, while any order will yield the same final estimate, a sensible convention is to process the cohorts in descending order of size, as this can enhance numerical stability.

Overall, this study provides a practical and scalable solution for valid inference in decentralized health data networks. The proposed sequential framework integrates well with modern machine learning-based nuisance estimation, supports doubly robust inference, and extends naturally to other outcome types and model classes. Future work will focus on generalizing these methods to semi-parametric and partially linear disease models with relaxed assumptions, enhancing their utility in real-world biomedical research.

## Data Availability

Michigan Genomics Initiative Data are available after institutional review board approval to select researchers. See at
All of Us' dataset is stored on the Researcher Workbench, a secure, cloud-based platform. Registered researchers can access data. See at

https://precisionhealth.umich.edu/our-research/michigangenomics/.

https://allofus.nih.gov/

## Acknowledgments

This work was supported through grant DMS1712933 from the National Science Foundation and MI-CARES grant 1UG3CA267907 from the National Cancer Institute. Data collection adhered to the Declaration of Helsinki principles. The University of Michigan Medical School Institutional Review Board reviewed and approved the consent forms and protocols of MGI study participants (IRB ID HUM00099605 and HUM00155849). Opt-in written informed consent was obtained. The authors acknowledge the Michigan Genomics Initiative participants, Precision Health at the University of Michigan, the University of Michigan Medical School Central Biorepository, the University of Michigan Medical School Data Office for Clinical and Translational Research, and the University of Michigan Advanced Genomics Core for providing data and specimen storage, management, processing, and distribution services, and the Center for Statistical Genetics in the Department of Biostatistics at the School of Public Health for genotype data curation, imputation, and management in support of the research reported in this work.

## Supplementary Materials

### S1 Variance Estimation

#### S1.1 CPL

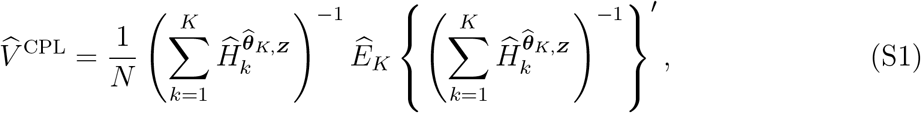

where the components are defined as follows:

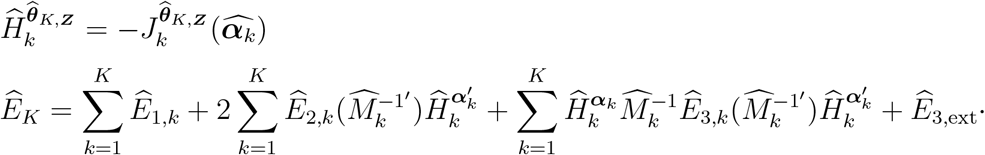

The individual components are defined as:

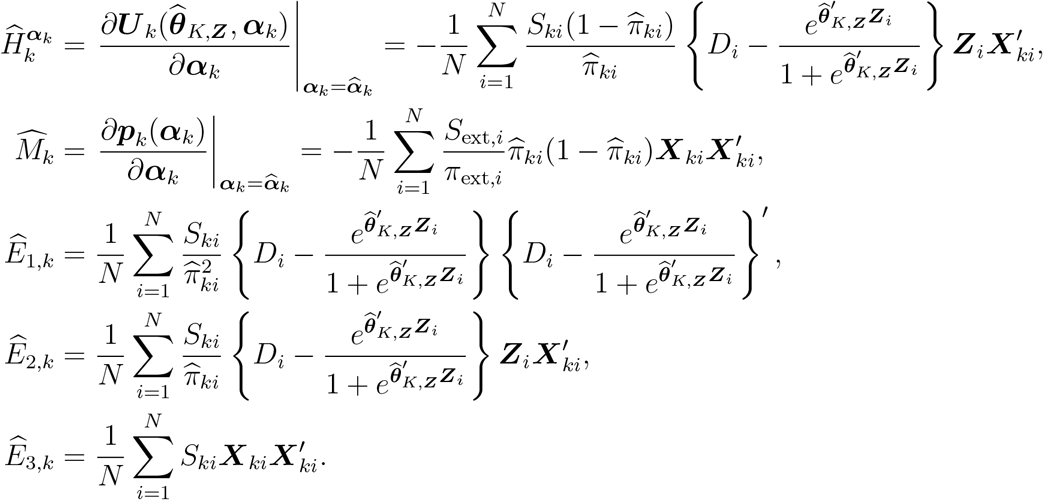

The external data correction term *Ê*_3,ext_ is:

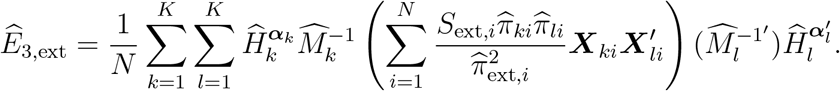

#### S1.2 SPL

For SPL, we use the same variance formula as in equation (S1), but compute it sequentially. For each cohort *k* = 1, 2, …, *K*, we evaluate the cohort-specific components 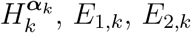, and *E*_3,*k*_ at the plug-in estimates 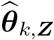 and 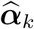.From the external data, we compute 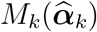 at each site and the cross-covariance term *E*_3,ext_ at the final site.

#### S1.3 CAIPW

We define the following notations:

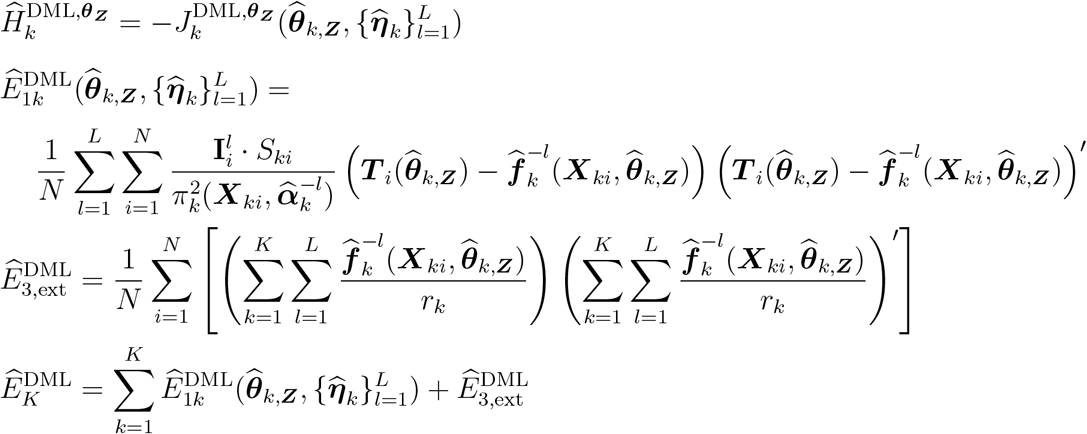

Finally we obtain the DML variance estimate by

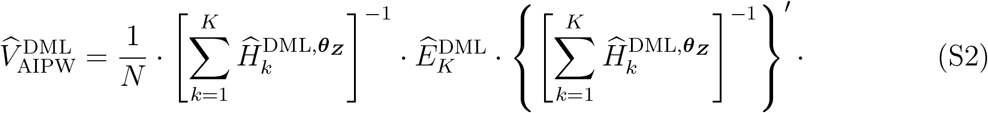

However, relying on this approach introduces bias in variance estimation if either the selection or the auxiliary model is misspecified, even when the point estimator remains consistent. This limitation is also discussed in the original work by Chernozhukov et al. (2018).

#### S1.4 SAIPW

For the SAIPW method, we adopt the same influence-function-based variance estimator as in CAIPW. However, in the sequential setting, the necessary quantities are computed sequentially across sites, consistent with the renewable estimation paradigm. At each site *k*, after estimating the site-specific parameter 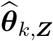 and the corresponding nuisance functions 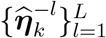, we compute the local contributions to the gradient matrix 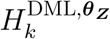 and the variance component of the influence function, denoted 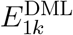. The external dataset is used to evaluate the cross-site covariance component 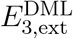, which is computed once and shared across sites. These components are cumulatively aggregated to obtain the final SAIPW variance estimator, which retains the same functional form as that given in Equation (S2).

### S2 Cancer Phenotypes List

In this study, phenotypes are defined using the PheWAS R package (Version 0.99.5-4), which maps ICD-9-CM and ICD-10-CM codes to PheWAS codes (PheCodes), resulting in up to 1,817 distinct codes. For our analysis, the PheCodes for cancer include: 145, 145.1, 145.2, 145.3, 145.4, 145.5, 149, 149.1, 149.2, 149.3, 149.4, 149.5, 149.9, 150, 151, 153, 153.2, 153.3, 155, 155.1, 157, 158, 159, 159.2, 159.3, 159.4, 164, 165, 165.1, 170, 170.1, 170.2, 172.1, 172.11, 174, 174.1, 174.11, 174.2, 174.3, 175, 180, 180.1, 180.3, 182, 184, 184.1, 184.11, 184.2, 185, 187, 187.1, 187.2, 187.8, 189, 189.1, 189.11, 189.12, 189.2, 189.21, 189.4, 190, 191, 191.1, 191.11, 193, 194, 195, 195.1, 195.3, 196, 197, 198, 198.1, 198.2, 198.3, 198.4, 198.5, 198.6, 198.7, 199, 199.4, 200, 200.1, 201, 202, 202.2, 202.21, 202.22, 202.23, 202.24, 204, 204.1, 204.11, 204.12, 204.2, 204.21, 204.22, 204.3, 204.4, 209, and 860.

**Table S1:** Simulation results comparing collaborative (CUW, CPL), meta (MUW, MPL), and joint (JUW, JPL) estimators. Metrics reported include relative bias percentage (RBP), root mean squared error (RMSE), relative efficiency (RE), and coverage probability (CP) for each target parameter *θ*_1_, *θ*_2_, and *θ*_3_.

| Method | RBP ($\theta_1$) | RBP ($\theta_2$) | RBP ($\theta_3$) | RMSE ($\theta_1$) | RMSE ($\theta_2$) | RMSE ($\theta_3$) | RE ($\theta_1$) | RE ($\theta_2$) | RE ($\theta_3$) | CP ($\theta_1$) | CP ($\theta_2$) | CP ($\theta_3$) |
| --- | --- | --- | --- | --- | --- | --- | --- | --- | --- | --- | --- | --- |
| CUW | 12.05% | 22.43% | 77.65% | 0.78 | 1.00 | 0.95 | 0.78 | 1.00 | 0.95 | 0.90 | 0.64 | 0.19 |
| MUW | 14.54% | 24.21% | 80.61% | 0.96 | 1.06 | 1.01 | 0.96 | 1.06 | 1.01 | 0.88 | 0.65 | 0.18 |
| JUW | 15.22% | 23.41% | 80.02% | 1.00 | 1.00 | 1.00 | 1.00 | 1.00 | 1.00 | 0.86 | 0.65 | 0.14 |
| CPL | 2.24% | 2.99% | 6.33% | 1.79 | 0.92 | 0.42 | 0.95 | 0.92 | 1.00 | 0.93 | 0.95 | 0.94 |
| MPL | 10.11% | 29.04% | 89.16% | 1.62 | 1.79 | 1.46 | 0.86 | 1.78 | 3.49 | 0.84 | 0.60 | 0.31 |
| JPL | 2.76% | 5.57% | 5.80% | 1.88 | 1.01 | 0.42 | 1.00 | 1.00 | 1.00 | 0.93 | 0.91 | 0.92 |

## References

1. J. Adler-Milstein and A. K. Jha. Hitech act drove large gains in hospital electronic health record adoption. Health affairs, 36(8):1416–1422, 2017.

2. All Of Us Research Programs Investigators. The “all of us” research program. New England Journal of Medicine, 381(7):668–676, 2019.

3. L. J. Beesley and B. Mukherjee. Statistical inference for association studies using electronic health records: handling both selection bias and outcome misclassification. Biometrics, 78 (1):214–226, 2022.

4. D. Blumenthal. Wiring the health system—origins and provisions of a new federal program. New England Journal of Medicine, 365(24):2323–2329, 2011.

5. D. Blumenthal and M. Tavenner. The “meaningful use” regulation for electronic health records. New England Journal of Medicine, 363(6):501–504, 2010.

6. Y. Chen, P. Li, and C. Wu. Doubly robust inference with nonprobability survey samples. Journal of the American Statistical Association, 115(532):2011–2021, 2020.

7. V. Chernozhukov, D. Chetverikov, M. Demirer, E. Duflo, C. Hansen, W. Newey, and J. Robins. Double/debiased machine learning for treatment and structural parameters, 2018.

8. R. Duan, M. R. Boland, J. H. Moore, and Y. Chen. Odal: A one-shot distributed algorithm to perform logistic regressions on electronic health records data from multiple clinical sites. In Pacific Symposium on Biocomputing. Pacific Symposium on Biocomputing, volume 24, page 30, 2019.

9. R. Duan, M. R. Boland, Z. Liu, Y. Liu, H. H. Chang, H. Xu, H. Chu, C. H. Schmid, C. B. Forrest, J. H. Holmes, et al. Learning from electronic health records across multiple sites: A communication-efficient and privacy-preserving distributed algorithm. Journal of the American Medical Informatics Association, 27(3):376–385, 2020.

10. R. Duan, Y. Ning, and Y. Chen. Heterogeneity-aware and communication-efficient distributed statistical inference. Biometrika, 109(1):67–83, 2022.

11. O. Efthimiou. Practical guide to the meta-analysis of rare events. BMJ Ment Health, 21(2):72–76, 2018.

12. M. Hu, X. Shi, and P. X.-K. Song. Collaborative inference for treatment effect with distributed data-sharing management in multicenter studies. Statistics in Medicine, 2024.

13. M. I. Jordan, J. D. Lee, and Y. Yang. Communication-efficient distributed statistical inference. Journal of the American Statistical Association, 2019.

14. R. Kundu, X. Shi, J. Morrison, J. Barrett, and B. Mukherjee. A framework for understanding selection bias in real-world healthcare data. Journal of the Royal Statistical Society Series A: Statistics in Society, 187(3):606–635, 2024a.

15. R. Kundu, X. Shi, M. Salvatore, L. G. Fritsche, and B. Mukherjee. A doubly robust method to counteract outcome-dependent selection bias in multi-cohort ehr studies. arXiv preprint 2412.00228, 2024b.

16. D.-Y. Lin and D. Zeng. On the relative efficiency of using summary statistics versus individual-level data in meta-analysis. Biometrika, 97(2):321–332, 2010.

17. Z. Liu, D. Wang, and Y. Pan. Superpopulation model inference for non probability samples under informative sampling with high-dimensional data. Communications in Statistics-Theory and Methods, 54(5):1370–1390, 2025.

18. L. Luo and P. X.-K. Song. Renewable estimation and incremental inference in generalized linear models with streaming data sets. Journal of the Royal Statistical Society Series B: Statistical Methodology, 82(1):69–97, 2020.

19. L. Luo, M. Risk, and X. Shi. Online causal inference with application to near real-time post-market vaccine safety surveillance. Statistics in Medicine, 2024.

20. K. Rice, J. P. Higgins, and T. Lumley. A re-evaluation of fixed effect (s) meta-analysis. Journal of the Royal Statistical Society Series A: Statistics in Society, 181(1):205–227, 2018.

21. M. Salvatore, R. Kundu, X. Shi, C. R. Friese, S. Lee, L. G. Fritsche, A. M. Mondul, D. Hanauer, C. L. Pearce, and B. Mukherjee. To weight or not to weight? the effect of selection bias in 3 large electronic health record-linked biobanks and recommendations for practice. Journal of the American Medical Informatics Association, page ocae098, 2024.

22. G. Schwarzer et al. meta: An r package for meta-analysis. R news, 7(3):40–45, 2007.

23. M. Zawistowski, L. G. Fritsche, A. Pandit, B. Vanderwerff, S. Patil, E. M. Schmidt, P. Vande-Haar, C. J. Willer, C. M. Brummett, S. Kheterpal, et al. The michigan genomics initiative: a biobank linking genotypes and electronic clinical records in michigan medicine patients. Cell Genomics, 3(2), 2023.

24. W. Zhou, M. Kanai, K.-H. H. Wu, H. Rasheed, K. Tsuo, J. B. Hirbo, Y. Wang, A. Bhattacharya, H. Zhao, S. Namba, et al. Global biobank meta-analysis initiative: Powering genetic discovery across human disease. Cell genomics, 2(10), 2022.

